# Analysis of Genetic Host Response Risk Factors in Severe COVID-19 Patients

**DOI:** 10.1101/2020.06.17.20134015

**Authors:** Krystyna Taylor, Sayoni Das, Matthew Pearson, James Kozubek, Marcin Pawlowski, Claus Erik Jensen, Zbigniew Skowron, Gert Lykke Møller, Mark Strivens, Steve Gardner

**Affiliations:** PrecisionLife Ltd, Long Hanborough, Oxford, UK

## Abstract

**BACKGROUND:** Epidemiological studies indicate that as many as 20% of individuals who test positive for COVID-19 develop severe symptoms that can require hospitalization. These symptoms include low platelet count, severe hypoxia, increased inflammatory cytokines and reduced glomerular filtration rate. Additionally, severe COVID-19 is associated with several chronic co-morbidities, including cardiovascular disease, hypertension and type 2 diabetes mellitus.

The identification of genetic risk factors that impact differential host responses to SARS-CoV-2, resulting in the development of severe COVID-19, is important in gaining greater understanding into the biological mechanisms underpinning life-threatening responses to the virus. These insights could be used in the identification of high-risk individuals and for the development of treatment strategies for these patients.

**METHODS:** As of June 6, 2020, there were 976 patients who tested positive for COVID-19 and were hospitalized, indicating they had a severe response to SARS-CoV-2. To overcome the limited number of patients with a mild form of COVID-19, we used similar control criteria to our previous study looking at shared genetic risk factors between severe COVID-19 and sepsis, selecting controls who had not developed sepsis despite having maximum co-morbidity risk and exposure to sepsis-causing pathogens.

**RESULTS:** Using a combinatorial (high-order epistasis) analysis approach, we identified 68 protein-coding genes that were highly associated with severe COVID-19. At the time of analysis, nine of these genes have been linked to differential response to viral pathogens including SARS-CoV-2. We also found many novel targets that are involved in key biological pathways associated with the development of severe COVID-19, including production of pro-inflammatory cytokines, endothelial cell dysfunction, lipid droplets, neurodegeneration and viral susceptibility factors.

**CONCLUSION:** The variants we found in genes relating to immune response pathways and cytokine production cascades, were in equal proportions across all severe COVID-19 patients, regardless of their co-morbidities. This suggests that such variants are not associated with any specific co-morbidity, but are common amongst patients who develop severe COVID-19. This is consistent with being able to find and validate severe disease biomarker signatures when larger patient datasets become available.

Several of the genes identified relate to lipid programming, beta-catenin and protein kinase C signalling. These processes converge in a central pathway involved in plasma membrane repair, clotting and wound healing. This pathway is largely driven by Ca^2+^ activation, which is a known serum biomarker associated with severe COVID-19 and ARDS. This suggests that aberrant calcium ion signalling may be responsible for driving severe COVID-19 responses in patients with variants in genes that regulate the expression and activity of this ion. We intend to perform further analyses to confirm this hypothesis.

Among the 68 severe COVID-19 risk-associated genes, we found several druggable protein targets and pathways. Nine are targeted by drugs that have reached at least Phase I clinical trials, and a further eight have active chemical starting points for novel drug development.

Several of these targets were particularly enriched in specific co-morbidities, providing insights into shared pathological mechanisms underlying both the development of severe COVID-19, ARDS and these predisposing co-morbidities. We can use these insights to identify patients who are at greatest risk of contracting severe COVID-19 and develop targeted therapeutic strategies for them, with the aim of improving disease burden and survival rates.

## Introduction

Coronavirus disease 2019 (COVID-19), caused by severe acute respiratory syndrome coronavirus 2 (SARS-CoV-2), is a major threat to public health. As of 16^th^ June 2020, there are estimated to be over 8 million confirmed cases globally, resulting in approximately 437,000 deaths worldwide^1,2^. Although many who develop the disease present with only mild symptoms, reports from multiple international health systems have shown that up to 20% of individuals testing positive for COVID-19 go on to develop severe forms of the disease that may require hospitalization.

Significant associations of disease severity risk have been observed with epidemiological factors including age, sex, blood group and ethnicity in addition to co-morbidity with many common conditions such as cardiovascular disease, diabetes, hypertension and chronic pulmonary diseases including asthma^3^. It would be clinically useful to be able to identify the features that result in differential host responses, particularly those that predispose some patients to developing severe COVID-19. In the context of management of at-risk populations, prior to an effective and widely available vaccine, such insights could have great utility in developing new detection, protection and treatment strategies targeted specifically at these high-risk individuals.

In our previous paper, we explored the shared risk factors between sepsis (a major clinical feature in hospitalized COVID-19 patients) and severe COVID-19 disease^4^. We observed that 59% of hospitalized COVID-19 cases also have sepsis^5^, and that the two diseases share similar co-morbidity risks^6^. In that study we identified 70 risk-associated genes in a sepsis population and found significant overlap in genetic risk variants between sepsis patients and those hospitalized with severe COVID-19.

As more data from COVID-19 patients has become available in the UK Biobank^7,8^, we are now able to investigate the host response genetic risk factors directly, using genotype datasets from 779, 877 and 929 patients hospitalized with severe COVID-19 and comparing them against healthy controls. In this study, we have sought to identify the risk variants associated directly with severe COVID-19 patients, to gain insight of the underlying pathology and disease mechanisms in relation to this patient group.

## Methods

COVID-19 test records were downloaded from the UK Biobank (data releases 18 May, 26 May and 6 June, 2020)^7,8^. The 18 May 2020 dataset included 5,657 test records relating to 3,002 individuals in UK Biobank. Of these, 1,073 patients had at least one positive COVID-19 test record, including 818 patients who were hospitalized and 255 who tested positive but were not hospitalized. In accordance with guidance from UK Biobank, we classified those 818 patients who were hospitalized after testing positive with COVID-19 as having a severe form of the disease and the 255 who were not hospitalized as having the mild form of the disease.

The analysis was conducted as a case-control study. After quality control and removal of samples with missing data, we assigned as cases 779 patients (442 males, 337 females) who had been hospitalized with severe COVID-19. Of these severe cases, 62% had one or more of the most common co-morbidities associated with high severe COVID-19 risk (Figure 1).

**Figure 1:**
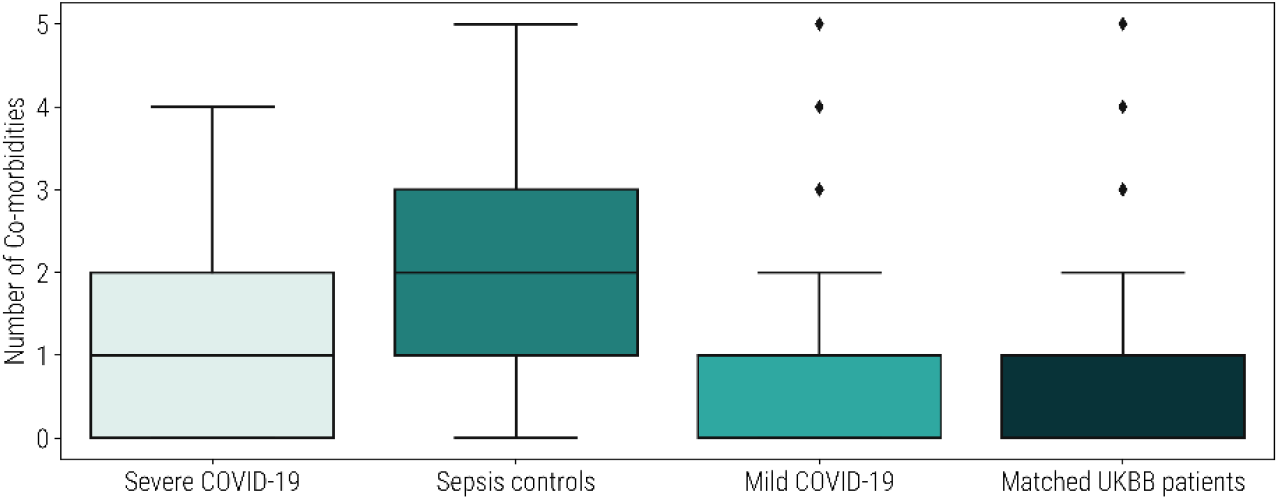
Boxplot showing the incidence of five co-morbidities associated with high COVID-19 risk (cardiovascular disease, hypertension, diabetes, chronic respiratory diseases and Alzheimer’s disease) among the May 18, 2020 severe COVID-19 patients, sepsis controls, mild COVID-19 patients and 77,900 randomly selected UK Biobank patients that were age and gender-matched with the severe COVID-19 patients.

The most prevalent co-morbidity was hypertension (50%), followed by chronic respiratory disease (22%), diabetes (20%) and cardiovascular disease (18%). 248 (32%) patients were reported having two or more co-morbidities. 291 severe COVID-19 cases had none of these co-morbidities. In comparison, these co-morbidities were found to be less prevalent in the mild cases and an age and gender-matched random selection of the UK Biobank population (Figure 1).

Due to the limited number of patients with confirmed mild disease we were unfortunately not able to use this cohort directly as our control set due to its small size and lack of statistical power. We therefore adopted similar control cohort criteria to our previous sepsis study^4^, based on evidence of shared risk factors and that severe COVID-19 patients often present with a similar pattern of co-morbid chronic conditions to those with sepsis. We selected patients who had not developed sepsis in spite of having been exposed to the most common sepsis-causing pathogens as well as having at least one of the most common chronic comorbidities known to increase a patient’s risk of developing sepsis and COVID-19 (Figure 2).

**Figure 2:**
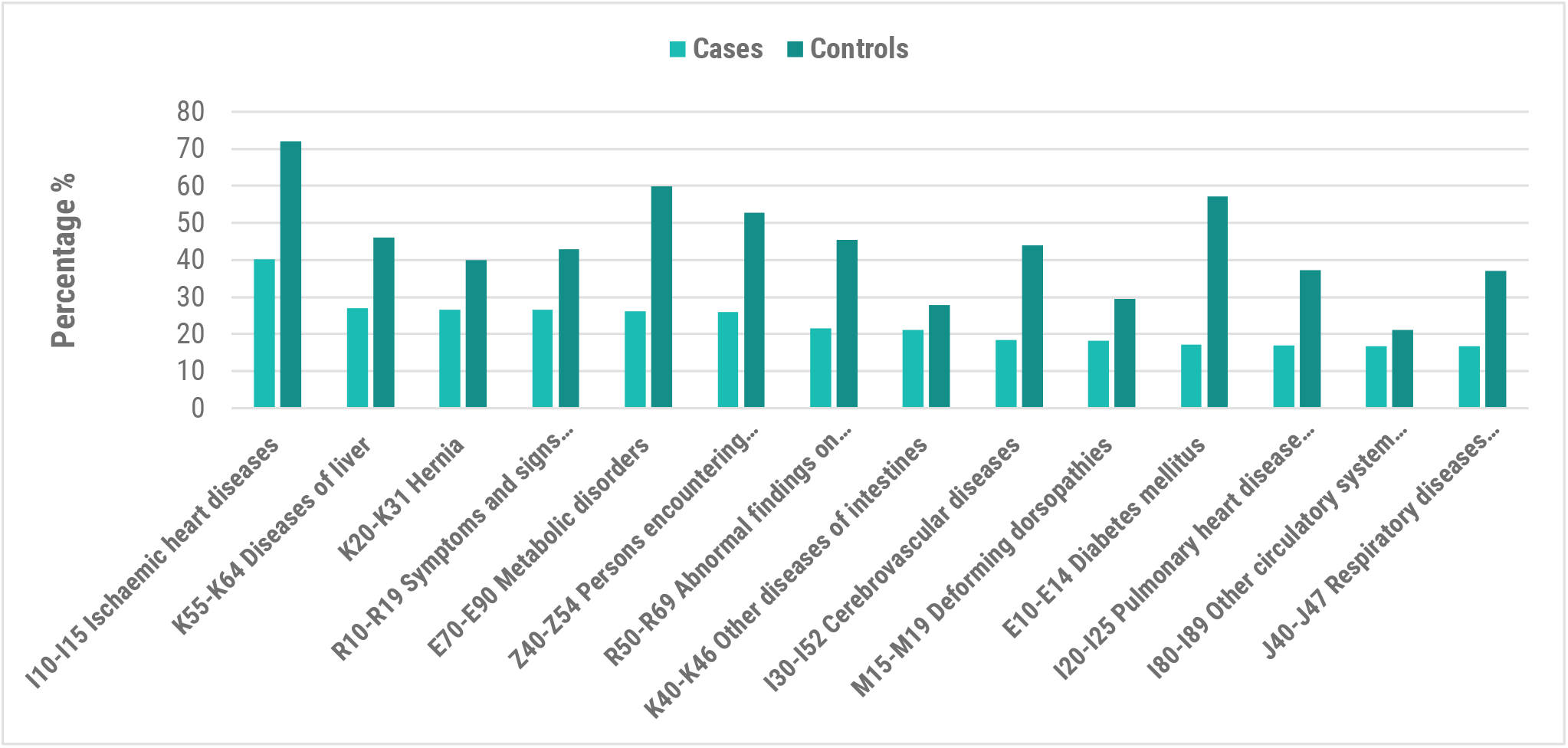
Incidence of co-morbidities by ICD-10 code from UK Biobank in the May 18, 2020 severe COVID-19 dataset (n= 2,332, 1:2 case control ratio with 779 cases and 1,553 controls).

We selected the oldest possible patients (as age is also a critical phenotypic risk factor for the severe form of COVID-19) with the highest number of co-morbidities. Controls were therefore selected for a lack of disease given maximum risk and exposure, and were gender-matched against the severe COVID-19 cases in the ratio 2:1. The exact control criteria and distribution of the co-morbidities in cases are described in the Appendix.

This set of patients represents the most similar available control cohort who could reasonably be expected to have some genetic protective effect against developing severe COVID-19 (or to lack any such risk factors). By selecting controls with a higher prevalence of similar chronic co-morbidities, we seek to ensure that the severe COVID-associated signal observed is not simply caused by the co-morbidities represented in the patient set but has the potential to be a true COVID-19 related enrichment.

After quality control (removal of SNPs with <95% coverage across subjects), the genotype data for the two cohorts contained 542,245 SNPs. The age distribution of cases and controls is shown in Figure 3.

**Figure 3:**
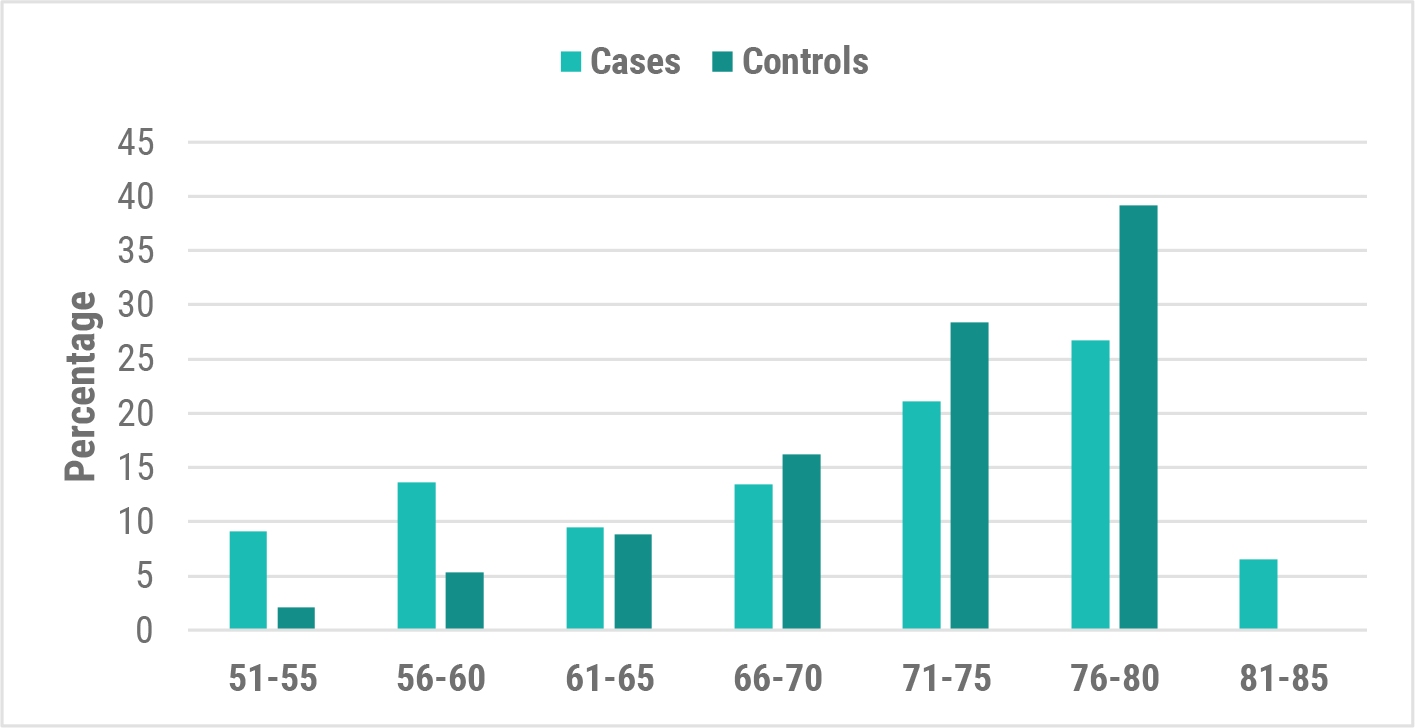
Age distribution of cases vs controls from UK Biobank in the May 18, 2020 severe COVID-19 dataset (779 cases and 1,553 controls).

We performed a SNP-based blood group analysis of the UK Biobank cohort by determining the blood groups (A, B, O and AB) of the cohort based on allele combinations of three SNPs (rs8176747, rs8176746, rs8176719) in the ABO gene^9^. The blood group frequencies of the cases and controls were calculated and two-sided Fisher’s exact tests were used to calculate blood-group specific odds ratios against the other blood groups (see Appendix).

Having generated our case-control dataset, we analyzed it using the **precision**life platform to identify risk-associated SNPs and genes that were strongly associated in the severe COVID-19 disease cohorts. This platform identifies high-order, disease-associated combinations of multi-modal (e.g. SNPs, transcriptomic, epidemiological or clinical) features at whole genome resolution in large patient cohorts. It has been validated across multiple different disease populations^10,11,12^. This type of analysis is intractable to other existing methods due the combinatorial explosion posed by the analysis of large numbers of patients with combinatorial non-linear additive combinations of features per patient.

When applied to genomic data, **precision**life finds high-order epistatic interactions (multi-SNP genotype signatures - typically of combinatorial order between 3-8) that are significantly more predictive of patients’ phenotype than those identified using existing single SNP based methods. When the individual SNPs making up these signatures are assessed individually across the whole population, they may fall below the GWAS (genome-wide association study) significance thresholds. However, we have demonstrated that when evaluated in combination with each other using multiple statistical validation techniques, these SNPs can be highly significant in particular disease sub-populations. The phenotype to which the signatures are associated in this context might be disease status, progression rate, therapy response or other, depending on the data available and study design.

We used the platform to find and statistically validate combinations of SNPs that together are strongly associated with the severe COVID-19 disease diagnosis. Analysis and annotation of these COVID-19-associated combinatorial genomic signatures took less than a day to complete, running on a dual CPU, 4 GPU compute server. The signatures identified by the analysis were then mapped to the human reference genome^13^ to identify disease-associated and clinically relevant target genes. A semantic knowledge graph derived from multiple public and private data sources was used to annotate the SNP and gene targets, including relevant tissue expression, chemical activity/tractability for gene targets, functional assignment and disease-associated literature. This provides contextual information to test the targets against the 5Rs criteria of early drug discovery^14^ and allows us efficiently to form strong, testable hypotheses for their mechanism of action in driving severe, life-threatening host responses to COVID-19 infection.

All of the significant disease signatures were traced back to the cases in which they were found and were associated with selected attributes such as case co-morbidities and COVID-19 test records. This generated a high-resolution stratification of severe COVID-19 patient subgroups and enabled further analysis of the different underlying factors relating to their specific forms of the disease.

Subsequently, UK Biobank added the COVID-19 test records of 1,508 individuals in data release 26 May, 2020 and 1,607 individuals in data release 6 June, 2020^8^. Of these, 401 patients were tested positive and 158 of them were hospitalized. We repeated our analysis on these two updated datasets with a higher number of controls to make the analysis more robust (see Appendix). 50 (∼70%) of the genes identified in our initial analysis were identified in more than one subsequent analysis (highlighted in bold in the gene tables) and therefore have an additional replication.

## Results

Using the severe COVID-19 dataset to perform a standard PLINK^15^ GWAS analysis revealed no significant SNPs (Figure 4) using a genome-wide significance threshold of *p*<5e-08. The lowest SNP PLINK *p*-value reported was 9.02e-08.

**Figure 4:**
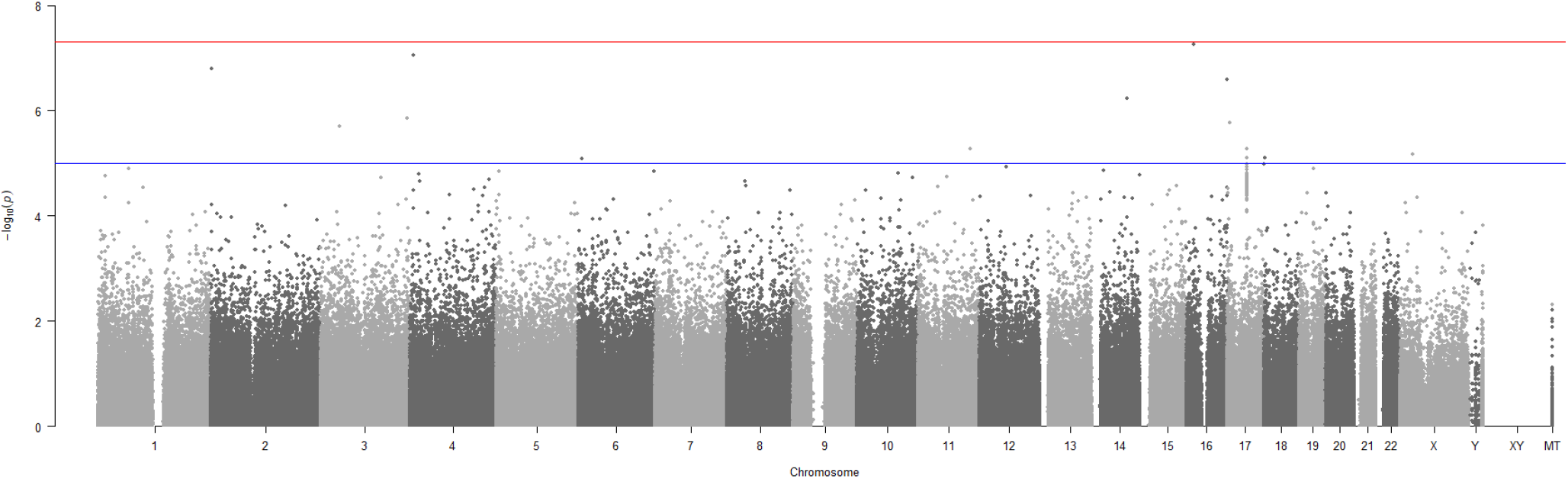
Manhattan plot generated using PLINK of genome-wide *p*-values of association for the May 18, 2020 severe COVID-19 UK Biobank cohort. The horizontal red and blue lines represent the genome-wide significance threshold at *p*<5e-08 and *p*<1e-05 respectively.

When the same May 18, 2020 severe COVID-19 patient dataset was run using the **precision**life platform, we identified 3,515 combinations of SNP genotypes representing different combinations of SNP genotypes that were highly associated with the severe COVID-19 patient cohort (Table 1). The majority (n = 3,494) of SNPs were found in combinations with 5 or more SNP genotypes, and as such could not have been found using standard GWAS analysis methods.

**Table 1:** Summary of May 18, 2020 Severe COVID-19 Cases vs Sepsis Controls disease study.

|  | Severe COVID19/Sepsis Study |
| --- | --- |
| **FDR** | 5% |
| **Disease signatures** | 3,515 |
| **SNPs in all Disease signatures** | 5,402 |
| **Penetrance**<br>(cases represented by all signatures) | 100% |
| **RF scored SNPs** | 156 |
| **RF scored Genes** | 71 |

All of the SNP genotypes and their combinations were scored using a Random Forest (RF) algorithm based on a 5-fold cross-validation method to evaluate the accuracy with which the SNP genotype combinations predict the observed case: control split. 156 SNPs were scored by the RF algorithm, indicating that they accurately predict the differences between cases and controls (Figure 5A). The chromosomal distribution of the critical SNPs is shown in aggregate in Figure 5B.

**Figure 5:**
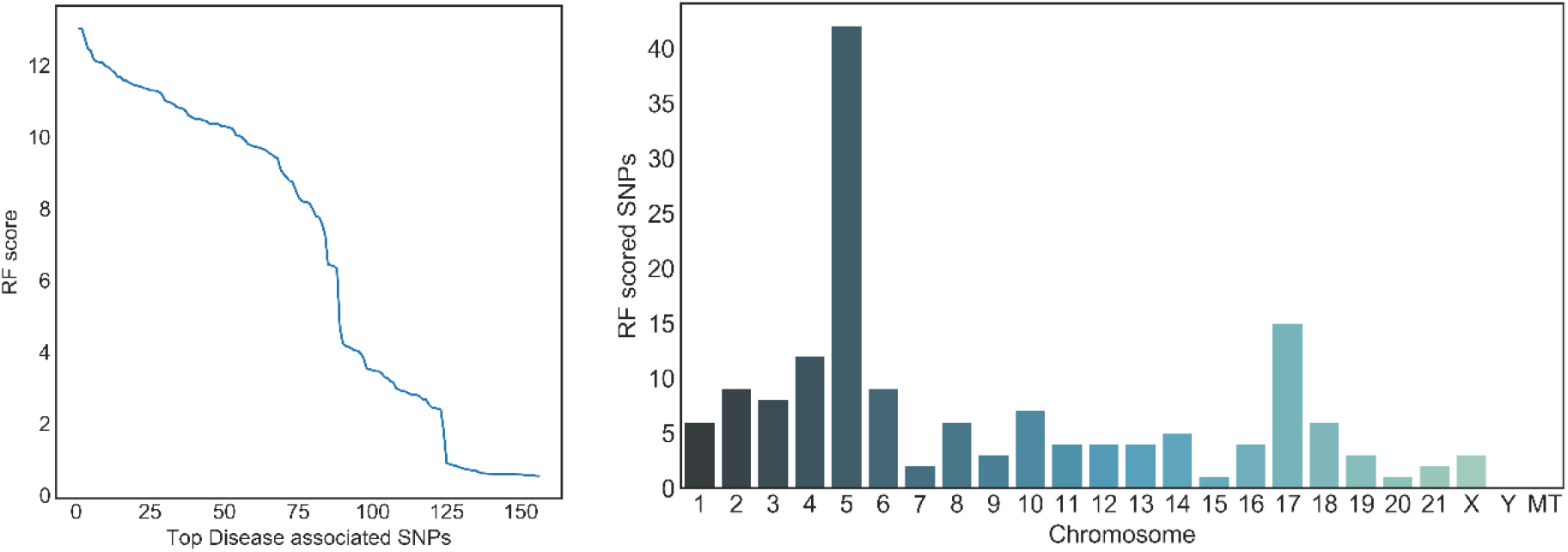
Distributions of (A) RF scores and (B) chromosomal locations for critical disease associated SNPs

Clustering the SNPs by the patients in whom they co-occur allows us to generate a disease architecture of severe COVID-19 patients, providing useful insights into patient stratification. We can use this to find genes and biological pathways that are associated with patient sub-populations and co-morbidities, enabling the development of disease biomarkers and precision medicine strategies.

Our analysis identified 68 protein-coding genes that were strongly associated with the disease phenotype in patients who developed severe COVID-19. To date, nine of these genes have already been linked to differential host responses to viral infections, including SARS-CoV-2 in various studies, providing validation for our hypothesis-free approach comparing severe COVID-19 patients against sepsis-free controls. We found several biological pathways and processes that were common in across the 68 COVID-associated genes, including T cell regulation and host pathogenic responses, inflammatory cytokine production, and lipid formation and endothelial cell function (Figure 9).

**Figure 6:**
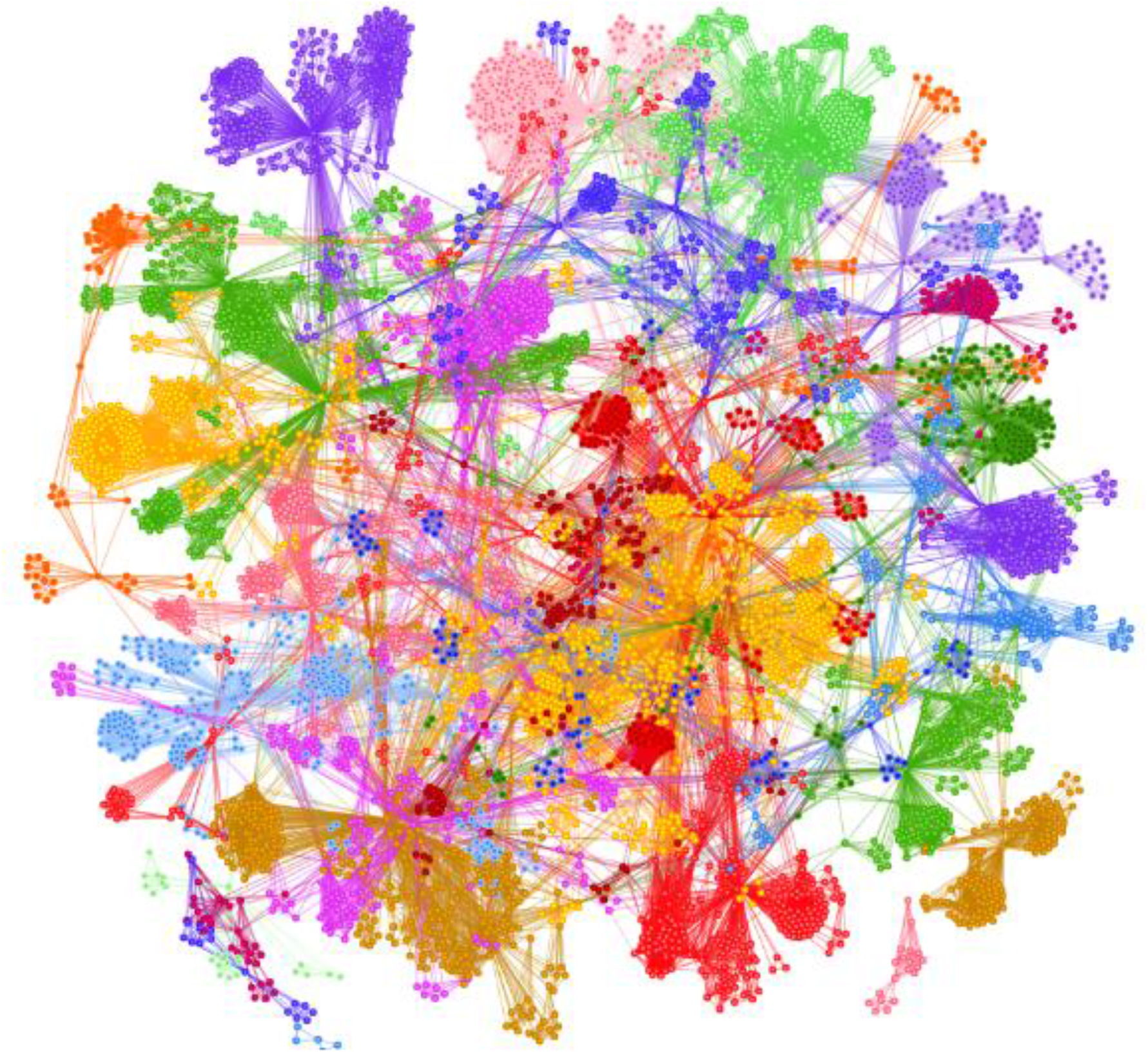
Disease architecture of the severe COVID-19 patient population generated by the **precision**life platform. Each circle represents a disease-associated SNP genotype, edges represent co-association in patients, and colours represent distinct patient sub-populations.

**Figure 7:**
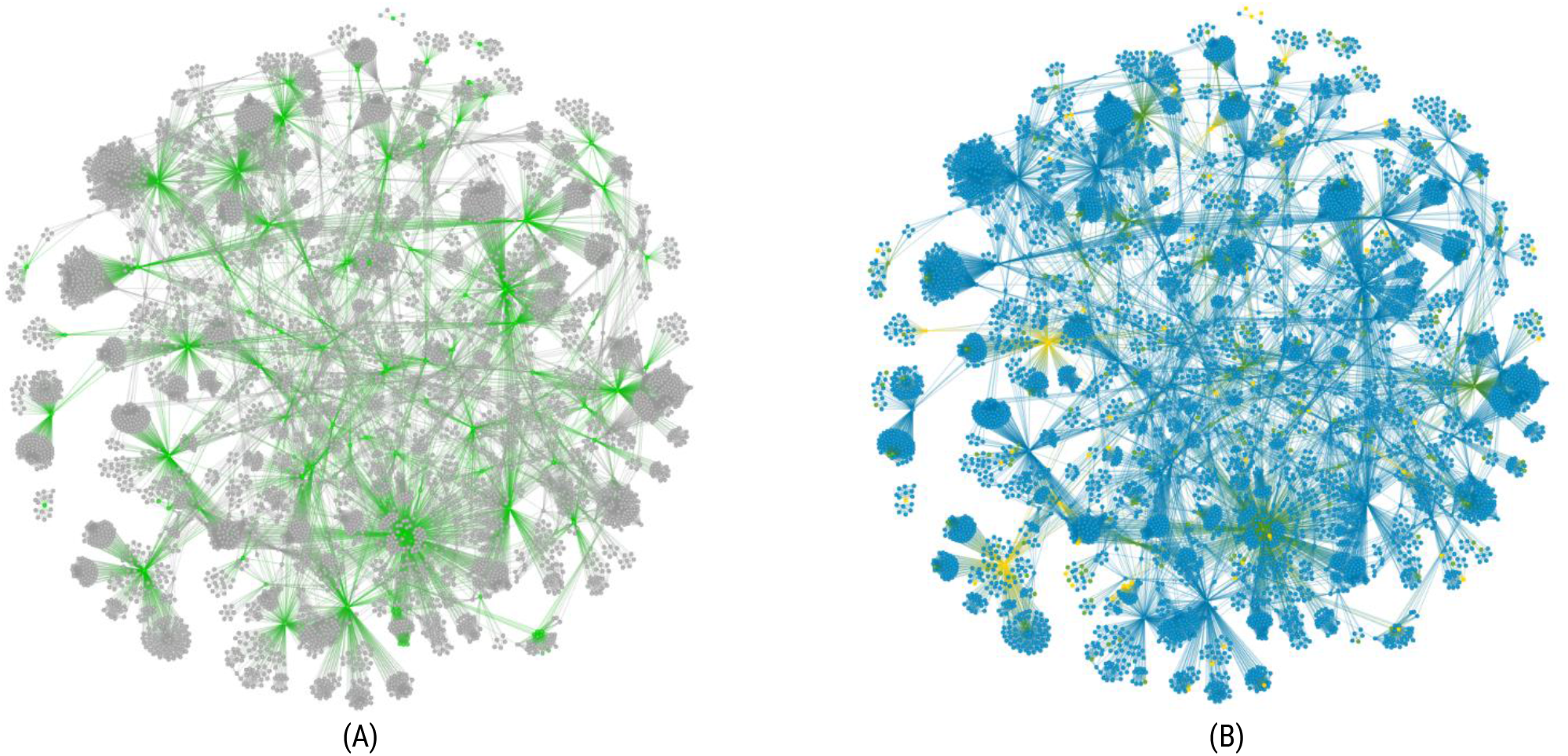
Disease architecture of the severe COVID-19 patient population highlighting (A) the critical disease SNPs (green) and (B) showing SNP genotypes (right - homozygous major allele = blue, heterozygous = green, homozygous minor allele = gold).

**Figure 8:**
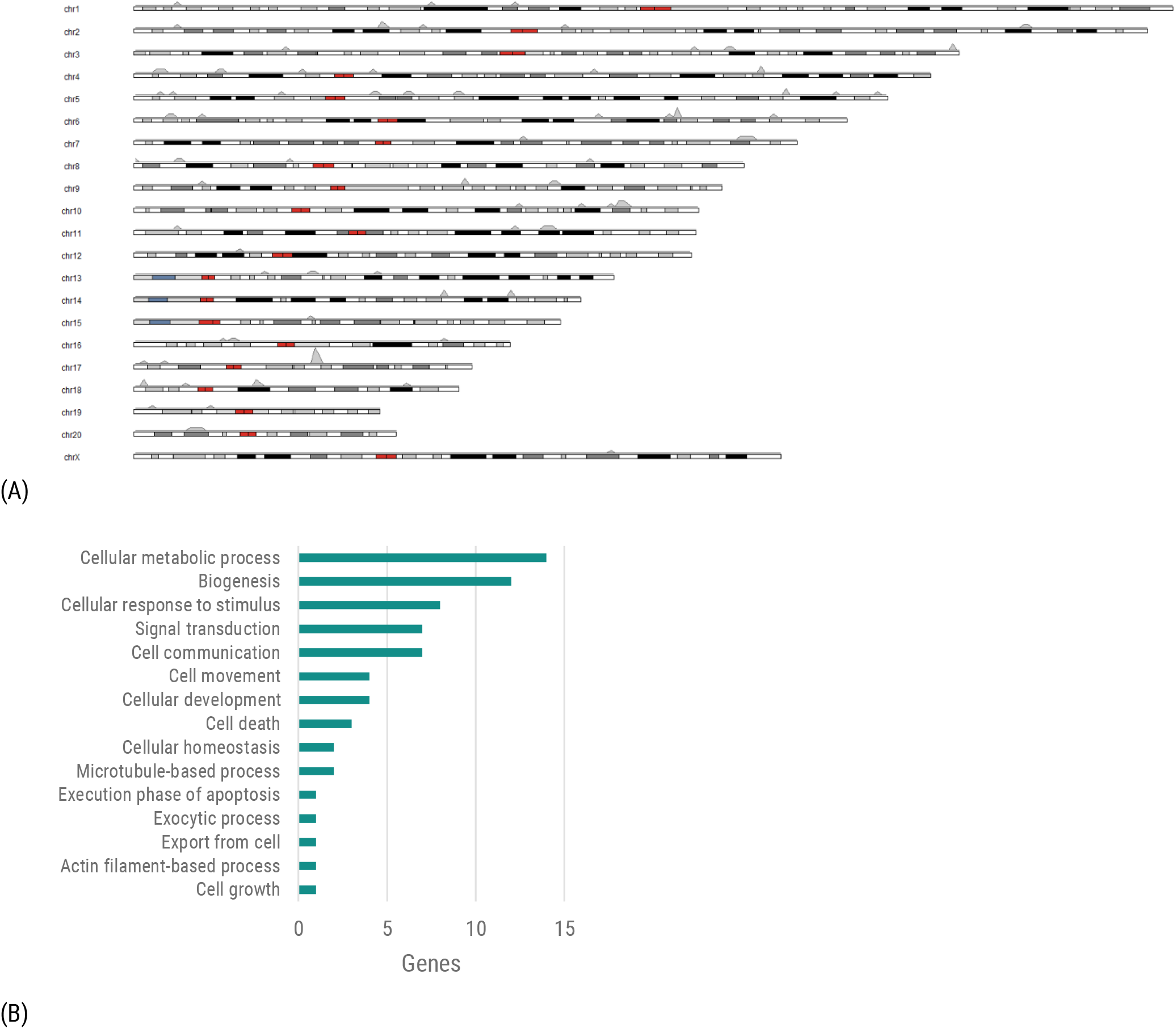
(A) Distributions of chromosomal locations for RF scored Genes. (B) Functional categories of genes defined by high-level Gene Ontology^16^ terms.

**Figure 9:**
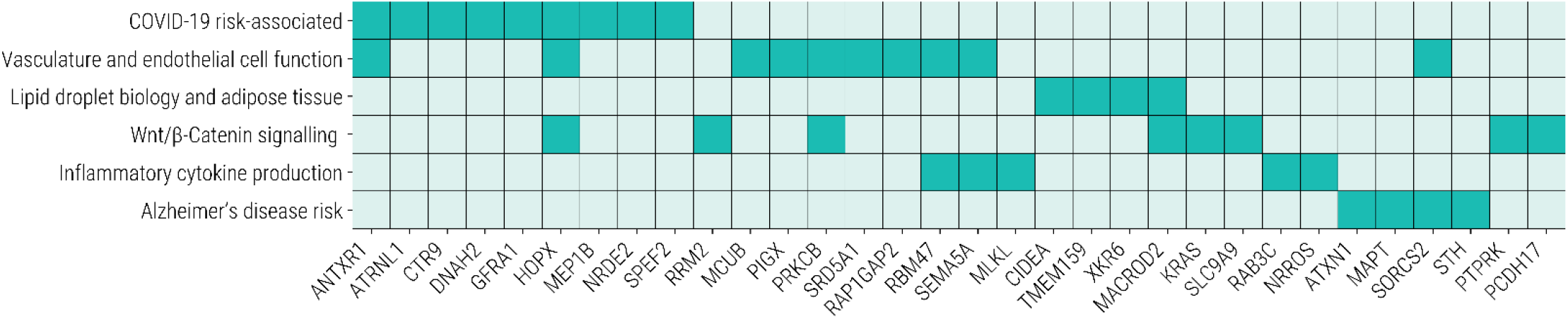
Biological pathways and processes known to be associated with some of the genes replicated across the datasets used in this study

We identified twelve genes that are associated with host immune response to viral infections, including SARS-CoV-2 (Table 2). Variants in several of these genes have been associated with increased infectivity to these strains. Genes marked in bold represent those that were further validated in subsequent analyses using the updated severe COVID-19 cases.

**Table 2:** Severe COVID-19 risk-associated genes associated with host immune response to virus infection, listed alphabetically. Bold text indicates genes found in multiple study populations.

| GENE | FUNCTION | MECHANISM OF ACTION |
| --- | --- | --- |
| **ANTXR1**<sup>17,18</sup> | Anthrax toxin receptor 1 (TEM8), type I transmembrane protein | ANTXR1 is used by anthrax toxin and Seneca valley virus (SVV) to gain entry into host cells |
| **ATRNL1**<sup>19</sup> | Attractin-like 1 | Phenotypes associated with decreased *Salmonella* invasion, hepatitis C virus replication and vaccinia virus infection |
| **CTR9** | Paf/RNA polymerase II complex component | Acts as a host restriction factor, suppressing HIV replication |
| **DNAH220** | Dynein heavy chain | Expressed in lung cilia, may be responsible for effective pathogenic clearance |
| **GFRA1**<sup>21</sup> | GDNF family receptor alpha | SNPs associated with susceptibility to hepatitis C and *Staphylococcus aureus* infection |
| **HOPX** | Hop homeodomain protein, cofactor | High expression in expanded CD8+ T cells, improving SARS-CoV-2 viral clearance |
| **IKZF2** | Ikaros family member, Helios | Helios expression is high in regulatory T cells, necessary for self-tolerance and prevention of autoimmunity |
| **ITK** | IL-2 inducible T cell kinase, Th2 differentiation | Altered expression associated with SARS-CoV-2 infectivity |
| **MEP1B** | Meprin A subunit beta, membrane metalloproteinase | Macrophages in *Mep1b*<sup>-/-</sup> mice have greater phagocytic function. Biomarker for high tuberculosis burden. |
| **NRDE2**<sup>22</sup> | RNA interference | RNA interference is a mechanism utilized by the innate immune response. NRDE2 is used by Kaposi's sarcoma-associated herpesvirus (KSHV) for late gene expression |
| **SPEF2**<sup>23</sup> | Sperm flagellar 2 | Loss of *Spef2* in mice results in increased inflammatory response to *Streptococcus pneumoniae* infection due to defects in pulmonary cilia. |
| **TBC1D2** | TBC1 domain family member, regulates Rab7A activation | SNP associated with severe H1N1 infection |

Our analysis also revealed five genes regulating pro-inflammatory pathways such as necroptosis, reactive oxygen species (ROS) production and cytokine signalling (Table 3).

**Table 3:**
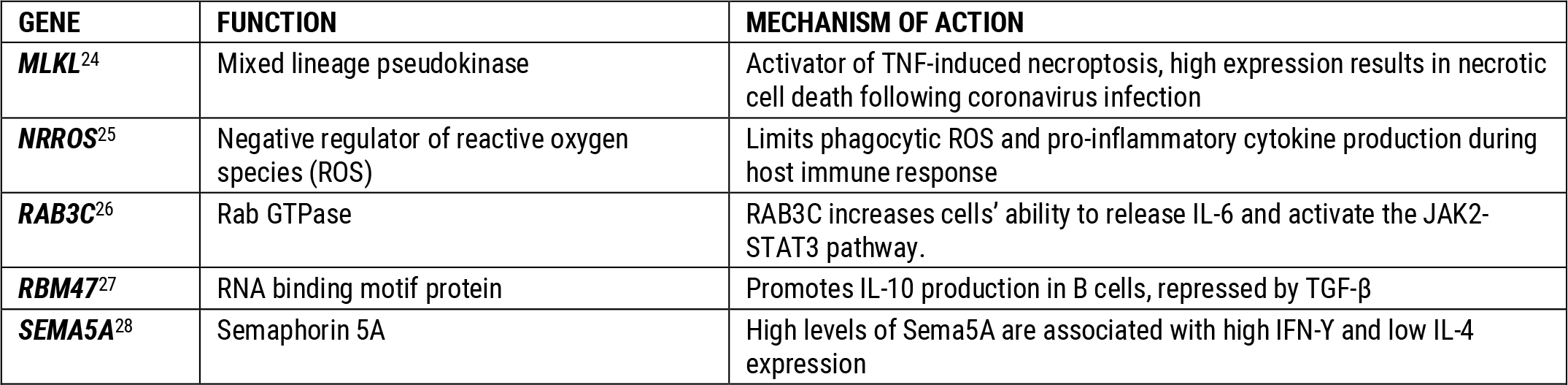
Severe COVID-19 risk-associated genes that have a role in regulation of inflammatory cytokine production

There were also six genes associated with severe COVID-19 patients that play central roles in lipid droplet biology, as well as having high expression in adipose tissue, correlating with serum lipid levels and coronary artery disease (Table 4).

**Table 4:** Severe COVID-19 risk-associated genes that are highly expressed in adipose tissue and relate to lipid storage and signalling

| GENE | FUNCTION | MECHANISM OF ACTION |
| --- | --- | --- |
| *CIDEA*<sup>29</sup> | Cell death inducing DFFA like effector | Co-localizes with perilipins, highly expressed in brown adipose tissue |
| *MACROD230* | Mono-ADP ribosylhydrolase | Highly associated with VAP-1 in human adipose tissue |
| *PLIN4*<sup>31</sup> | Perilipin 4 | Lipid droplet protein, highly expressed in white adipose tissue, high expression results in cardiac steatosis |
| *SLIT3*<sup>32</sup> | Slit guidance ligand 3 | High expression in adipose tissue controlled by *Prdm16* expression. Upregulated in obese patients. |
| *TMEM159*<sup>33,34</sup> | Promethin | Activated by peroxisome proliferator-activated receptor γ1 (PPARγ1), partners with seipin to regulate lipid droplet organization |
| *XKR6*<sup>35</sup> | XK-related 6 | SNP association with serum lipid levels and risk of CAD |

In addition, our analysis revealed 12 more genes that may be implicated in the vascular complications seen in COVID-19 (Table 5). These are all highly expressed in the cardiovascular system, modulating cardiac function and endothelial cell homeostasis.

**Table 5:** Severe COVID-19 risk-associated genes that are implicated in cardiovascular and endothelial cell function

| GENE | FUNCTION | MECHANISM OF ACTION |
| --- | --- | --- |
| *ANTXR1*<sup>36</sup> | Anthrax toxin receptor 1, type I transmembrane protein (TEM8) | Loss of ANTXR1 results in endothelial basement membrane loss and leaky blood vessels |
| *HOPX*<sup>37</sup> | Hop homeodomain protein, cofactor | Acts via serum response factor (SRF), modulating the expression of cardiac genes and stress-induced cardiac hypertrophy |
| *MCUB*<sup>38</sup> | Mitochondrial calcium uniporter | Role in cardiac homeostasis, response to cardiac stress including ischemia |
| *PIGX*<sup>39</sup> | Phosphatidylinositol glycan anchor biosynthesis class X | Associated with hypertension. Forms a complex with PIG-M that regulates several processes, including maintenance of arterial blood pressure. Acts via apelin receptor signalling. |
| *PRKCB*<sup>40</sup> | Protein kinase C beta form, protein kinase | Variety of different cellular functions, including endothelial cell proliferation and insulin signaling |
| *PLS3*<sup>41</sup> | Plastin-3 | Induced by angiotensin II in endothelial cells to promote cell migration |
| *RAP1GAP2*<sup>42,43</sup> | GTPase activator for RAP-1A | Regulates platelet granule secretion and aggregation |
| *RBM47*<sup>44</sup> | RNA binding motif protein | Missense variant is associated with hypertension |
| *SEMA5A*<sup>45</sup> | Semaphorin 5A | High *Sema5A* expression increased endothelial cell proliferation and angiogenesis and decreased apoptosis |
| *SLIT3*<sup>46</sup> | Slit guidance ligand 3 | Upregulation associated with endothelial cell dysfunction in pulmonary hypertension |
| *SORCS2*<sup>47</sup> | Sortilin-related VPS10 domain-containing receptor 2 (SorCS2), oxidative stress response gene | SorCS2 releases endostatin, an endogenous inhibitor of endothelial cell proliferation and angiogenesis |
| *SRD5A1*<sup>48</sup> | Steroid 5 alpha-reductase 1, catalyzes the production of androgen dihydrotestosterone | Androgens have been linked to peripheral artery disease and macrophage modulation |

Amongst other cancer-associated genes, we identified nine that directly interact with the Wnt/β-catenin signaling pathway (Table 6). Except for KRAS and SLC9A9, all of the proteins encoded by these genes act as endogenous inhibitors of the pathway.

**Table 6:**
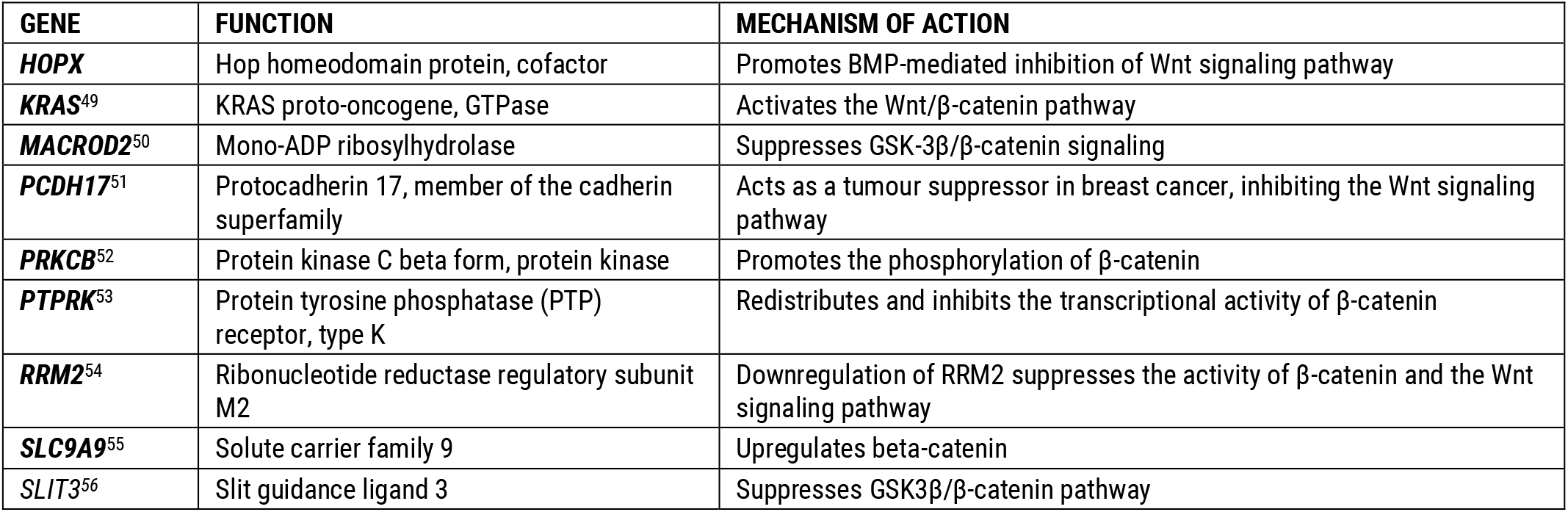
Severe COVID-19 risk-associated genes that directly interact with the Wnt/β-Catenin signalling pathway

Finally, our analysis identified four genes that have previously been associated with increased risk of developing Alzheimer’s disease (AD), including *MAPT*, the key gene underlying tau pathology (Table 7).

**Table 7:**
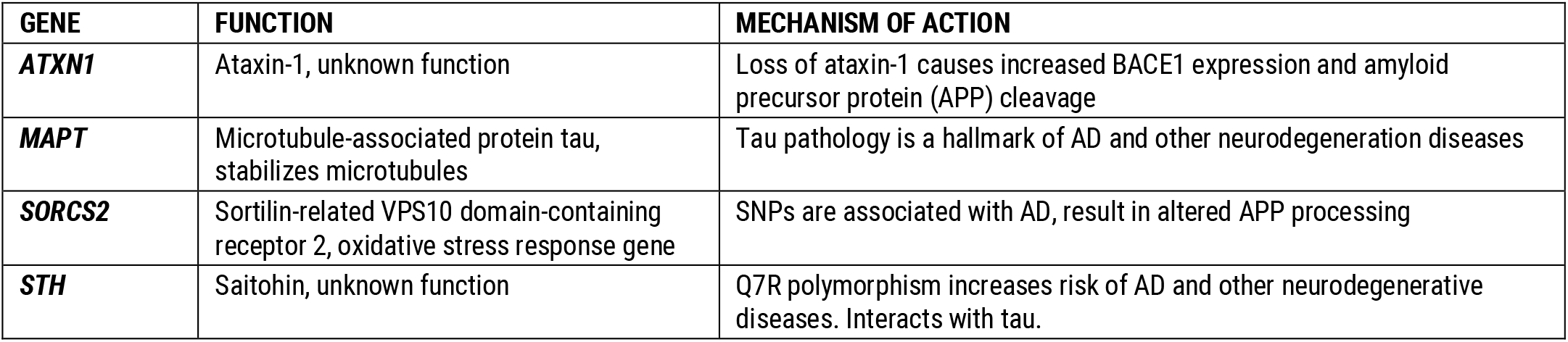
Severe COVID-19 risk-associated genes that have also been found to confer increased Alzheimer’s disease risk

| GENE | FUNCTION | MECHANISM OF ACTION |
| --- | --- | --- |
| *ATXN1* | Ataxin-1, unknown function | Loss of ataxin-1 causes increased BACE1 expression and amyloid precursor protein (APP) cleavage |
| *MAPT* | Microtubule-associated protein tau, stabilizes microtubules | Tau pathology is a hallmark of AD and other neurodegeneration diseases |
| *SORCS2* | Sortilin-related VPS10 domain-containing receptor 2, oxidative stress response gene | SNPs are associated with AD, result in altered APP processing |
| *STH* | Saitohin, unknown function | Q7R polymorphism increases risk of AD and other neurodegenerative diseases. Interacts with tau. |

**Table 8:** Pairwise comparison of co-morbidities prevalent in severe COVID-19 patient population

|  | All Severe COVID-19 | Cardiovascular Disease | Hypertension | Respiratory Disease | Diabetes |
| --- | --- | --- | --- | --- | --- |
| All Severe COVID-19 | 779 |  |  |  |  |
| Cardiovascular Disease | 136 |  |  |  |  |
| Hypertension | 385 | 115 |  |  |  |
| Respiratory Disease | 170 | 39 | 105 |  |  |
| Diabetes | 143 | 55 | 117 | 38 |  |
| Alzheimer's Disease | 10 | 1 | 5 | 1 | 2 |

**Table 9:**
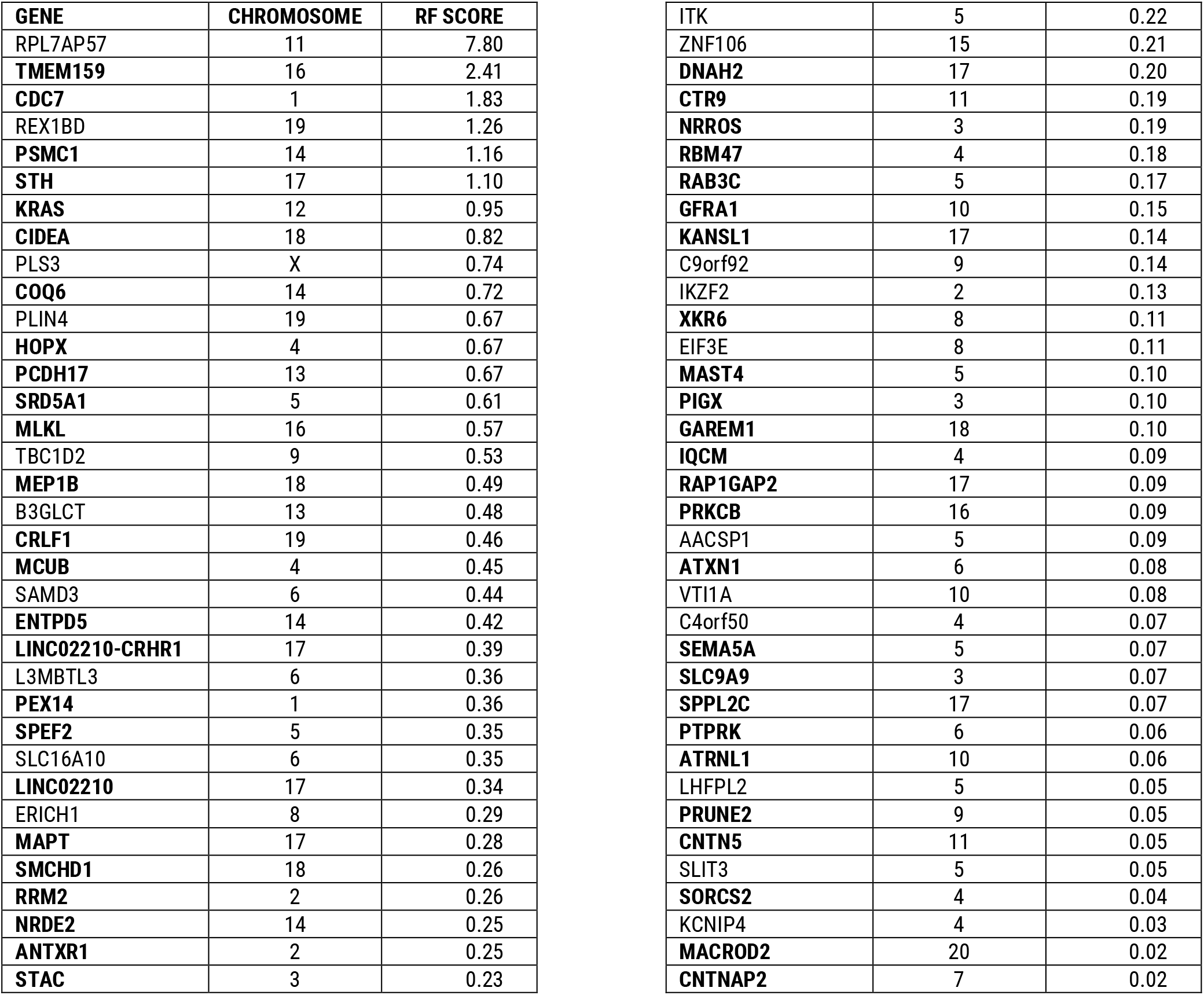
Table of severe COVID-19-associated genes with chromosomal location and RF score (genes in bold have been validated in further studies)

**Table 10.** Table showing drug repurposing candidates for 10 target genes identified as being COVID-19 related

| GENE | COMPOUND CHEMBL ID | DRUG | PHASE | MOLECULE TYPE |
| --- | --- | --- | --- | --- |
| CDC7 | CHEMBL3544943 | bms-863233 | 2 | Small molecule |
| CDC7 | CHEMBL3545090 | rxdx-103 | 1 | Small molecule |
| CDC7 | CHEMBL3545321 | nms-1116354 | 1 | Small molecule |
| PSMC1 | CHEMBL325041 | bortezomib | 4 | Small molecule |
| PSMC1 | CHEMBL451887 | carfilzomib | 4 | Protein |
| PSMC1 | CHEMBL2103884 | oprozomib | 1 | Small molecule |
| PSMC1 | CHEMBL3545432 | ixazomib citrate | 4 | Small molecule |
| SRD5A1 | CHEMBL254328 | abiraterone | 4 | Small molecule |
| SRD5A1 | CHEMBL1200969 | dutasteride | 4 | Small molecule |
| RRM2 | CHEMBL467 | hydroxyurea | 4 | Small molecule |
| RRM2 | CHEMBL1637 | gemcitabine hydrochloride | 4 | Small molecule |
| RRM2 | CHEMBL1750 | clofarabine | 4 | Small molecule |
| RRM2 | CHEMBL1096882 | fludarabine phosphate | 4 | Small molecule |
| RRM2 | CHEMBL1200983 | gallium nitrate | 4 | Small molecule |
| RRM2 | CHEMBL3544910 | motexafin gadolinium | 3 | Small molecule |
| RRM2 | CHEMBL3989496 | tezacitabine | 2 | Small molecule |
| ITK | CHEMBL1201733 | pazopanib hydrochloride | 4 | Small molecule |
| ITK | CHEMBL4085457 | pf-06651600 | 3 | Small molecule |
| ITK | CHEMBL1873475 | ibrutinib | 4 | Small molecule |
| CIDEA | CHEMBL121 | rosiglitazone | 4 | Small molecule |
| PLIN4 | CHEMBL121 | rosiglitazone | 4 | Small molecule |
| MLKL | CHEMBL3220918 | necrosulfanamide | preclinical | Small molecule |
| GFRA1 | CHEMBL2108380 | liatermin | 1 | Small molecule |
| PRKCB | CHEMBL300138 | enzastaurin | 3 | Small molecule |
| PRKCB | CHEMBL91829 | ruboxistaurin | 3 | Small molecule |
| PRKCB | CHEMBL494089 | gsk-690693 | 1 | Small molecule |
| PRKCB | CHEMBL574737 | ucn-01 | 2 | Small molecule |
| PRKCB | CHEMBL565612 | sotrastaurin | 2 | Small molecule |
| PRKCB | CHEMBL608533 | midostaurin | 4 | Small molecule |
| PRKCB | CHEMBL3545332 | cep-2563 | 1 | Small molecule |

**Table 11.** Table showing 8 target genes identified as being COVID-19 related that have active compounds in ChEMBL^56^

| GENE | NO. OF ACTIVE COMPOUNDS (ChEMBL) |
| --- | --- |
| MACROD2 | 3 |
| MAST4 | 6 |
| MLKL | 11 |
| SLC16A10 | 70 |
| MEP1B | 90 |
| KRAS | 120 |
| L3MBTL3 | 122 |
| MAPT | 9,589 |

**Table 12.** Table showing all the UK Biobank COVID-19 datasets and numbers of cases:controls (post-QC) used in this study

| UK Biobank Data Release | 18 May 2020 | 26 May 2020 | 6 June 2020 |
| --- | --- | --- | --- |
| **Cases** | 779<br>(442 males, 337 females) | 877<br>(492 males, 385 females) | 929<br>(524 males, 405 females) |
| **Controls** | 1,553 | 5,438 | 5,563 |

**Table 13.** Results of the ABO blood group analysis. Two-sided Fisher’s exact tests were used to calculate blood-group specific odds ratios against the other blood groups for the analyses. P-values <0.05 are shown in bold.

| Blood group analysis | Severe cases (779) vs Sepsis controls (1553) |  | Severe cases (779) vs all UK Biobank individuals (488,295) |  |
| --- | --- | --- | --- | --- |
|  | Odds Ratio | P-value | Odds Ratio | P-value |
| A vs AB/B/O | 1.201 | **0.037** | 1.120 | 0.096 |
| O vs A/B/AB | 0.880 | 0.154 | 0.860 | 0.050 |
| B vs A/B/O | 1.010 | 1.000 | 1.120 | 0.334 |
| AB vs A/B/O | 0.750 | 0.296 | 0.850 | 0.500 |

Out of the total 68 genes, nine are targeted by clinical candidates that have been evaluated in Phase I clinical trials and beyond (see Appendix). These could potentially form the basis of repurposing therapies, after evaluating factors such as safety and pharmacology data. A further eight targets have active chemistry in ChEMBL^57^, meaning they have active chemical starting points for novel drug development (see Appendix). We can also extend our search for repurposing candidates by looking into known gene interactions to find other more tractable targets in implicated in the same biological pathway.

## Discussion

Grouping the 68 genes by common biological functions revealed that many are involved in processes that have also been linked to aberrant host responses to COVID-19, such as the pro-inflammatory cytokine storm and immune system dysregulation, as well as lipid droplet formation and endothelial cell dysfunction.

### ABO BLOOD-GROUP ASSOCIATION

We found that blood group A was significantly overrepresented in the cases compared to the sepsis-free controls (p=0.037) and all UK Biobank individuals (p=0.096), indicating that it confers a risk factor for developing severe COVID-19. On the other hand, blood group O was found in fewer severe COVID-19 cases (p=0.154) than the controls and the UK Biobank as a whole (p=0.05). This supports the findings that having O blood group conferred a protective effect against developing respiratory failure during COVID-19^58^.

### HOST RESPONSE FACTORS AND INFLAMMATION

The host immune response must maintain a balance between effective viral clearance and limiting the immune response to prevent chronic inflammation and collateral tissue damage. In many patients who develop severe COVID-19 reactions and acute respiratory distress syndrome (ARDS) there is evidence of dysregulated cytokine production, resulting in increased levels of pro-inflammatory mediators including IL-1, IL-6, IL-8, CXCL-10^59^, causing pathological features such as inflammatory cell infiltration, pulmonary edema and sepsis^60^.

From the number of genes we found relating to immune response pathways and cytokine production cascades, it is clear that patients who develop severe COVID-19 and ARDS may have innate genetic variants that prevent this balance from being struck. These variants were found in equal proportions across all severe COVID-19 patients, regardless of their co-morbidities. This suggests that variants in these genes may not be associated with any specific co-morbidity, but are common amongst many patients who develop severe COVID-19.

HOPX regulates a variety of different cellular processes, including cardiac development and myogenesis^37^. However, it was reported in a recent COVID-19 study as part of a selection of genes upregulated in expanded CD8+ T cells in patients with mild COVID-19^61^. These expanded CD8+ effector T cells likely represent SARS-CoV-2-specific T cells, indicating greater efficiency in viral clearance in those patients. However, it is also necessary for Th-1 persistence and resistance to apoptosis, driving chronic inflammation and autoimmune mechanisms^62^. This exemplifies the delicate balance between establishing an effective host immune response necessary for viral clearance and preventing the development of chronic inflammation that results in collateral tissue damage. It seems likely that patients who develop severe COVID-19 may have an inherent imbalance in these factors.

Similarly, we identified SNP variants in *ITK* that were highly associated with severe COVID-19. High *ITK* expression is associated with Th2-driven diseases such as allergic asthma, causing pro-inflammatory cytokine production, eosinophil infiltration and production of mucus^63^. A range of potent and selective inhibitors (both small molecule and biologic) of ITK have been developed by several pharmaceutical companies^64,65^. No selective ITK inhibitors have progressed beyond preclinical testing to date, although ibrutinib – a joint ITK and BTK inhibitor – is currently licensed for use in B cell malignancies and as also demonstrated efficacy in the treatment of leishmaniasis. Data collected from several clinical trials indicate that ibrutinib is reasonably well-tolerated by patients^66^. Inhibition of ITK diminishes lung injury, cytokine production and inflammation in a mouse model of asthma^67^. Many of the pro-inflammatory cytokines, such as IFNγ, IL-2, and IL-17, blocked by ITK inhibition are raised in patients with ARDS^68,69^. Furthermore, use of a selective ITK inhibitor blocked HIV cell entry, transcription and particle formation, effectively reducing viral replication^70^.

A study has found that MLKL is implicated in necrotic cell death following infection with a neurovirulent human coronavirus (HCoV)^71^. Whilst MLKL-induced necrosis may be useful in limiting viral replication, increased necroptosis can also lead to increased inflammation and tissue damage. SNP variants in *MLKL* could be predisposing patients to severe COVID-19 in one of two ways; with under-functioning protein resulting in poor viral clearance, or through over-expression and over-activation of the necroptotic inflammatory response resulting in organ damage. The SNP variant identified in *MLKL* was found in the highest scoring 20% of the significant genes in the severe COVID-19 dataset and is present in 251 cases. It is not highly associated with any of the most common co-morbidities found in this dataset. Necrosulfonamide (NSA) is a specific inhibitor of MLKL that potently suppresses necroptosis^72^. Inhibition of this pathway using NSA decreases pro-inflammatory cytokines such as IL-1β, IL-6 and IL-17A in a way that was protective in a model of psoriasis. High levels of IL-1β and IL-6 have both been observed in cases with severe COVID-19^73^. Unfortunately, NSA has only ever been evaluated in preclinical trials, so there is no safety or toxicology data in humans available.

NRROS (LRRC3) is an inhibitor of multiple toll-like receptors (TLR) and subsequent NF-ĸB signalling, acting as a brake on pro-inflammatory cytokine production^74^. NRROS also limits the amount of reactive oxygen species (ROS) produced by phagocytes during the innate immune response, thereby limiting tissue damage caused whilst defending against invading pathogens. It could therefore be hypothesized that SNP variants in this gene, limiting its activity, could make individuals more susceptible to COVID-19-induced inflammatory damage^75^. This is supported by the evidence that *Lrrc33-/-* mice suffered from increased organ damage as a result of greater pro-inflammatory signalling when challenged with LPS. There are currently no specific small molecule activators of NRROS in publicly-available databases, however a number of chemical agents have been shown to increase the expression of both the protein and mRNA forms of this gene which may provide chemical starting points for novel drug discovery^76^.

RAB3C may be driving inflammation by inducing the release of IL-6 and activation of the IL-6/JAK2/STAT3 pathway, driving the production of pro-inflammatory cytokines^26,77^. Ruxolitinib – a JAK2 inhibitor – has been shown to mitigate the effects of RAB3C-induced IL-6 release^25^. Ruxolitinib is currently being trialled as a treatment for respiratory distress caused by SARS-CoV-2^78^.

### VASCULAR INFLAMMATION, AUTOPHAGY AND CARDIOVASCULAR DYSFUNCTION

Endothelial dysfunction and vascular inflammation, resulting in neutrophilic infiltration, endothelial cell apoptosis and tissue edema, is seen in patients who develop ARDS from SARS-CoV-2 infection^79^. Many of the common co-morbidities associated with severe COVID-19 development, - such as diabetes, hypertension and cardiovascular disease – are already associated with vascular inflammation and endothelial cell dysfunction pathological features^80,81,82^. It is therefore unsurprising that these patients are at higher risk of developing severe COVID-19.

Many of the risk-associated SNPs that we found that were related to these pathological pathways were found in severe COVID-19 patients who had at least one of these co-morbidities. However, we controlled for this effect by co-morbidity-matching these cases against the sepsis-free controls, meaning that these SNPs are not likely to be just be artefacts of the co-associated co-morbidity, but a differentiating factor in the development of severe COVID-19 and ARDS.

Helios (*IZKF2*) expression is used as a marker for regulatory T cells (Tregs), and therefore has an important role in self-tolerance and regulating autoimmunity^83^. Decreased levels of Helios+ Tregs have been observed in patients with hypertension and rheumatoid arthritis, with lower expression levels correlating with increased inflammatory markers^84^. Tregs may protect against hypertension by limiting vascular inflammation through suppression of effector T cells^85,86^. Furthermore, our analysis revealed that variants in *IZKF2* were disproportionately co-associated with patients with hypertension (Figure 10). This adds further evidence to the association between vascular inflammation, severe COVID-19 and cardiovascular hypertensive co-morbidities.

**Figure 10:**
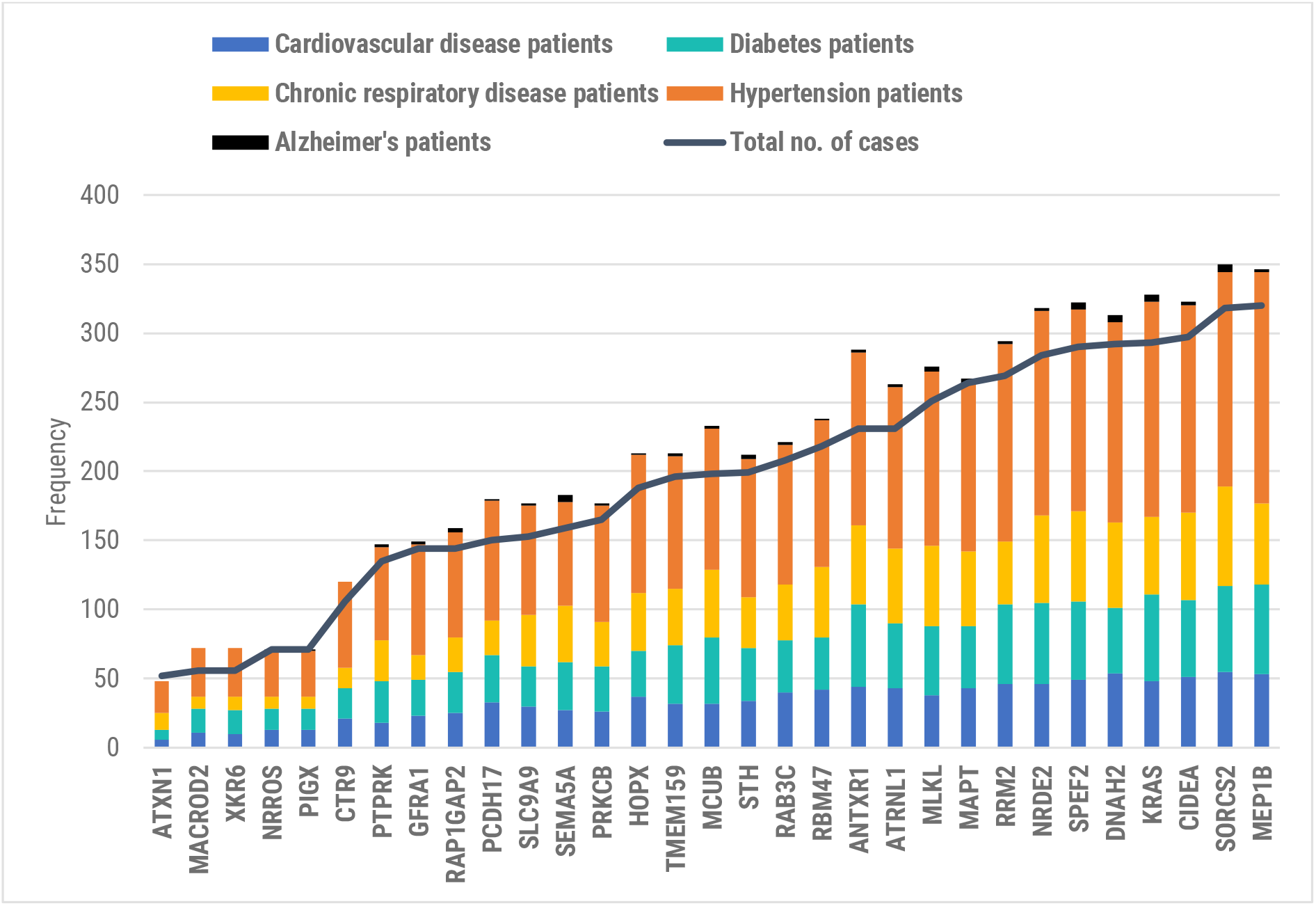
Stacked bar plots showing the number of cases with different co-morbidities (cardiovascular disease, hypertension, diabetes, chronic respiratory diseases and Alzheimer’s disease) associated with the severe form of COVID-19 who are affected by the risk-associated genes identified by the **precision**life platform. The line plot shows the total unique number of cases who are affected for each gene.

*MCUB* encodes one of the pore-forming subunits of the mitochondrial Ca^2+^ uniporter (MCU). MCUB is a necessary part of a protective response against mitochondrial Ca^2+^ overload during cardiac injury and ischemia^87^. *Mcub -/-* mice displayed increased cardiac remodelling and ischemic injury as a result of increased mitochondrial Ca^2+^ uptake^88^. Although more research is required to fully understand the role of MCUB in severe COVID-19 pathogenesis, a decreased level of serum calcium has been suggested as a biomarker for increased COVID-19 severity and ARDS^89^.

PKCβ (*PRKCB*) is a serine/threonine protein kinase that is activated by calcium. It has a range of functions, including B cell activation, endothelial cell proliferation and activation of apoptosis^40^. However, PKCβ has also been linked to a number of different vascular diseases, including atherosclerosis, diabetes and hypertension^90,91^. We find that in our case population, the risk-associated SNP found in *PRKCB* was found in 165 severe COVID-19 cases (penetrance = 20.2%), and was present in 45% of patients with cardiovascular disease and 51% of patients with hypertension. High levels of PKCβ result in increased vascular inflammation, endothelial dysfunction and oxidative stress, all of which have been found in patients with severe COVID-19^92,93^.

PKCβ also drives the accumulation of cholesterol in macrophages, leading to foam cell development and macrophage dysregulation^94^. Therefore, inhibition of PKCβ could help to reverse some of the vascular-related pathology, contributing to sepsis development and multi-organ failure, that is seen in severe COVID-19 patients. Ruboxistaurin (LY333531) is a selective PKCβ inhibitor that has been trialled as an anti-diabetic drug to reduce vascular and retinal complications that certain diabetic patients develop^95^. It has reasonable pharmacokinetic and toxicology data, as it can be orally administered and is well-tolerated by patients^96,97^. However, it was discontinued and has failed to progress beyond Phase III clinical trials^98^.

We also found genes, including *CIDEA* and *PLIN4*, that protect against insulin resistance (IR). IR results in increased cardiovascular inflammation and oxidative stress, contributing to atherosclerotic plaque formation and hypertension^99^. High Cidea expression decreases circulating fatty acid levels by increasing the level of triglycerides stored in lipid droplets. This helps to protect against insulin resistance^100^. The anti-diabetic drug rosiglitazone may have insulin sensitising effects by increasing the expression of both Cidea and perilipins^29^. Our analysis identified *CIDEA* and a perilipin, *PLIN4*, as being highly associated with severe COVID-19 patients. SNPs in *CIDEA* were found in one of the highest proportions in severe COVID-19 patients, particularly in patients with cardiovascular and hypertensive diseases (Figure 10). This indicates that insulin resistance is a contributing factor to vascular inflammatory pathologies that may predispose patients to developing ARDS in response to SARS-CoV-2.

Finally, variants in *GFRA1* are within the highest 10% of genes most associated with hypertensive patients in our case population. GFRα1, encoded by *GFRA1*, plays a key role in glial cell line-derived neurotrophic factor (GDNF)-mediated signalling^101^. GFRα1 induces autophagy via activation of the RET/AMPK signalling pathway independently from its interaction with GDNF. In the early stages of sepsis, autophagy is a protective mechanism employed by cells to remove damaged proteins, reduce mitochondrial dysfunction-induced inflammation and eliminate pathogens^102,103^. Furthermore, inhibition of autophagy in mouse models of sepsis results in increased mortality. Selective agonists of GFRα1 are under development^104^, which may help to protect against sepsis-induced tissue damage in the early stages of the disease.

### LIPID DROPLET BIOLOGY

There is increasing evidence that lipid dysfunction plays a role in COVID-19 pathogenesis. Lipid droplets (LD) play a key role in viral pathogenesis, as intracellular lipid stores are crucial for viral replication and assembly^105^. LD proteins have been associated with particle load and pathogenicity in other virus strains, including hepatitis C, dengue and rotavirus^106^. It has also been shown that coronaviruses use host cellular lipid machinery during replication^107^.

We identified six genes associated with lipids and adipose tissue, with three genes directly involved in lipid droplet formation (Table 4). A different member of the perilipin family (PLIN3) is used by hepatitis C virus (HCV) for steatosis development and viral assembly^108^, with PLIN3 inhibition resulting in decreased viral particle release from host cells^109^. Promethin (*TMEM159*) has also been recently identified as lipid droplet organiser (LDO) in partnership with seipin^34^. Although promethin has not been studied in the context of viral disease, overexpression of its co-associated protein seipin results in significantly reduced viral particle secretion and infectivity in a model of hepatitis C^110^.

### WNT/β-CATENIN SIGNALLING PATHWAY

Our analysis identified nine genes that regulate either the Wnt or β-catenin components of the Wnt/β-catenin pathway.

There is a long-established role of the Wnt gene-family and the associated signally pathway in embryogenesis and therefore its relationship to developmental disorders and carcinogenesis. However recent work^111,112^ shows an emerging and complex picture of the role of Wnt genes in host cell defence mechanisms, the modulation of inflammatory cytokine production and connections between innate and adaptive immune systems. The role of these ligands varies within the family members and their relation to β-catenin.

Wnt/β-catenin signalling has both anti- and pro-inflammatory effects in different contexts, dependent on its interaction with NF-ĸB^111^. In several studies, activation of β-catenin was shown to inhibit IL-1β and the subsequent production of IL-6 and matrix metalloproteinases (MMPs), resulting in an anti-inflammatory effect^112,113^. Although its role in sepsis pathogenesis is yet to be fully elucidated, patients with severe sepsis and sepsis-driven ARDS had increased levels of Wnt5A in lung tissue and serum^114^.

However, in bronchial epithelial cells, inhibition of beta-catenin’s activation of NF-ĸB resulted in fewer proinflammatory cytokines and cell injury^115^. In a previous unpublished analysis using the same combinatorial approach on a Sjögren’s Syndrome cohort found in the UK Biobank, we identified a significant number of genes involved in the Wnt/β-catenin pathway. As with severe COVID-19, patients with Sjögren’s syndrome also present with elevated cytokines and inflammation-driving pathology^116^.

The Wnt/ β -catenin pathway has also been implicated in promoting viral replication. In a model of influenza (H1N1), activation of this pathway resulted in higher viral replication and inhibition of Wnt/ β -catenin signalling using iCRT14 reduced viral production and improved clinical symptoms^117^. It has been demonstrated that Rift Valley Virus exploits the proliferative cell state that activation of Wnt signalling promotes to enable viral replication through easier trafficking of viral proteins within the cell. It has also been shown that coronaviruses use this mechanism in proliferative cells^118^. Therefore, inhibition of the Wnt/β-catenin pathway may help limit SARS-CoV-2 replication and reduce viral load in this way. This theory is supported by the fact that niclosamide – an anti-helminthic drug – limits coronavirus replication in a model of SARS^119^. The study did not investigate how this drug inhibited viral replication, but a subsequent study has revealed that niclosamide is an inhibitor of the Wnt signalling pathway^120^. Due to the significant number of genes found to regulate this pathway in our analysis, we believe that modulation of this pathway could be of benefit to a large number of patients who develop severe COVID-19.

### CALCIUM SIGNALLING PATHWAY

Several of the processes mentioned above, including lipid programming, beta-catenin and protein kinase C signalling, converge in a central pathway involved in plasma membrane repair, clotting and wound healing^121^. This pathway is largely driven by Ca^2+^ activation, which is a known serum biomarker associated with severe COVID-19 and ARDS^89^. Of the 68 risk-associated genes found in our analysis, at least 16 of them having calcium-binding domains or are dependent on Ca^2+^ signalling.

Calcium (Ca^2+^) drives wound constriction and clotting via F-actin^121^. Beta-catenin binds to F-actin and plays a role in endothelial barrier function^122,123^. Calcium also plays a role in lipid patterning, altering the plasma membrane composition in response to cell injury. Finally, activation of diacylglycerol (DAG and protein kinase c (including the β isoform) results in vesicle replenishment and wound repair^124,125^.

The pathological observations seen in severe COVID-19 of thrombocytopaenia, leaky blood vessels and multi-nucleated fusion cells^126,127^, support the hypothesis that these Ca^2+^/lipid/F-actin pathways may converge into a pathological process that drives life-threatening reactions to COVID-19 such as sepsis and ARDS.

Furthermore, the gene cluster at 3p21.31 identified as a susceptibility locus for respiratory failure in COVID-19^58^ contains several genes that are involved in calcium ion signalling; *SLC6A29* is a regulator of calcium-dependent amino acid uptake, and *CXCR9* and *CCR9* act as a signalling molecules by increasing the level of intracellular Ca^2+^. This suggests that aberrant calcium ion signalling may be responsible for driving severe COVID-19 responses in patients with variants in genes that regulate the expression and activity of this ion.

We intend to perform further analyses to confirm this hypothesis.

### NEURODEGENERATIVE DISEASE ASSOCIATIONS

There is emerging evidence that key genes associated with Alzheimer’s Disease (AD) risk such as *APOE4* are associated with increased risk of severe COVID-19 disease, although the reasons for this link are still unclear^128^. Our analysis identified a strong conserved signal for four genes that also confer greater risk of developing AD, including *MAPT*.

We also found three additional genes that were significant in a previous neurodegenerative disease study we performed. This unpublished study identified several gene variants in which were highly associated in cases with sporadic amyotrophic lateral sclerosis (ALS). We have also shown that increasing the activity of two of these genes in a mixed cell neuronal assay containing SOD1 reactive astrocytes significantly improves motor neurone survival. This could indicate shared biological pathways that drive both severe COVID-19 and neurodegeneration, potentially through pro-inflammatory, neurotoxic mechanisms. However, the SNP variants found in these genes were not highly enriched in the few severe COVID-19 patients who had been diagnosed with AD. This is likely due to the relatively low numbers of cases diagnosed with AD in UK Biobank as a whole.

## Conclusion

We performed this analysis using only genetic data from patients found in the UK Biobank, identifying 68 protein coding genes that are highly associated with the development of severe COVID-19. These targets would not have been found using standard analytical approaches such as GWAS on the same population. In addition to this, we have identified 29 drugs and clinical candidates and a further eight targets with chemical starting points that could be used in the development of treatment strategies that improve clinical outcomes in severe COVID-19 patients.

It appears that the variants we found in genes relating to immune response pathways and cytokine production cascades were in equal proportions across all severe COVID-19 patients, regardless of their co-morbidities. This suggests that such variants are not associated with any specific co-morbidity, but are common amongst patients who develop severe COVID-19. In contrast there were small deviations in the penetrance of some of the SNPs identified in different co-morbidity cohorts. While the pattern of these deviations were similar across cardiovascular, diabetes and hypertension co-morbidity cohorts, the pattern across the respiratory co-morbidity cohort was somewhat different. These differences, while suggestive of the need for further study, were not yet significant enough to be reported in detail.

As more test records and additional medical data become available in the UK Biobank and other data sources, we will be able to fully ascertain the severe and mild COVID-19 cases and provide an additional layer of validation to the results from this study.

One limitation of the UK Biobank dataset is that the ethnicity distribution of the participants is heavily skewed to white British participants and it has consequently not been possible to fully investigate additional risk factors in BAME patients. We are actively seeking to investigate severe COVID-19 risk factors in other datasets with more ethnically diverse populations. The addition of more phenotypic and clinical data in our analyses may also be used to gain greater understanding into the association of other observed epidemiological risk factors such as ethnicity, socioeconomic status and prescription medication history with development of severe COVID-19.

## Data Availability

All data described in this paper will be made available in supplemental files and via the COVID-19 Knowledge Graph https://github.com/covid19kg and https://www.biorxiv.org/content/10.1101/2020.04.14.040667v1

## Acknowledgements

We would like to acknowledge UK Biobank for providing us access to the data under Application ID 44288 and Bugbank for linking infection data from Public Health England to improve the study of infection in the UK Biobank cohort.

## Appendix

### SEPSIS CONTROL CRITERIA

1. Controls to <u>exclude</u> any patients with the following ICD-10 codes:

2. Controls to include at least one of the following ICD-10 codes:

| ICD-10 | Disease |
| --- | --- |
| A02.1 | Salmonella septicemia |
| A22.7 | Anthrax septicemia |
| A40.x | Streptococcal septicemia |
| A41.x | Other septicemia |
| B37.7 | Candidal septicemia |
| O35 | Puerperal sepsis |
| R57.2 | Septic shock |

3. Controls to include least one of the following ICD-10 codes:

2. Controls to include at least one of the following ICD-10 codes:
| ICD-10 | Disease |
| --- | --- |
| J12-J18 | Pneumonia |
| J20-J22 | Lower respiratory infection |
| B95 | Streptococcus |
| B95 | Staphylococcus |

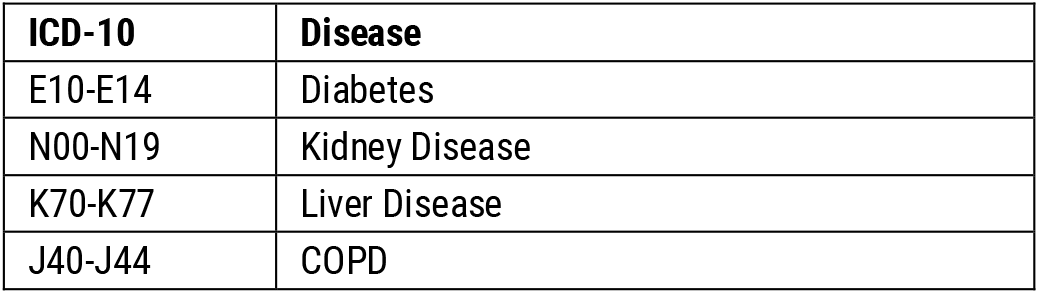

### IDENTIFICATION OF CASES WITH SPECIFIC CO-MORBIDITIES

Cases with co-morbidities were identified using the following disease codes:

IDENTIFICATION OF CASES WITH SPECIFIC CO-MORBIDITIES Cases with co-morbidities were identified using the following disease codes:
| Disease | ICD-10 | Self-Reported codes | Operation codes |
| --- | --- | --- | --- |
| Cardiovascular disease | I20-I25 | 1070, 1075, 1095 | K40-41, K45, K49, K50.2, K75 |
| Hypertension | I10, I11, R03 | 1065 |  |
| Diabetes | E10-14 | 1220, 1222, 1223 |  |
| Chronic respiratory diseases | J40-47 | 1111, 1112 |  |
| Alzheimer's disease | G30 |  |  |

